# New-onset chronic musculoskeletal pain following COVID-19 infection fulfil the Fibromyalgia clinical syndrome criteria

**DOI:** 10.1101/2024.07.10.24310234

**Authors:** Omar Khoja, Matthew Mulvey, Sarah Astill, Ai Lyn Tan, Manoj Sivan

**Affiliations:** Leeds Institute of Rheumatic and Musculoskeletal Medicine, University of Leeds, Leeds, UK; Academic Unit of Palliative Care, Leeds Institute of Health Sciences, University of Leeds, Leeds, UK; School of Biomedical Sciences, Faculty of Biological Sciences, University of Leeds, Leeds, UK; NIHR Leeds Biomedical Research Centre, Leeds Teaching Hospitals NHS Trust, Leeds, UK; COVID Rehabilitation Service, Leeds Community Healthcare NHS Trust, Leeds, UK

**Keywords:** Long COVID, Post-COVID-19 Condition, Post-Acute Sequela, FM syndrome, Chronic pain, Chronic widespread pain, Post-Acute Infection Syndrome (PAIS)

## Abstract

**Background:** New-onset chronic musculoskeletal (MSK) pain (> 3 months duration) is one of the commonest persistent symptoms of Post-COVID syndrome (PCS). There is emerging evidence that the chronic MSK pain and associated symptoms in PCS have similarities to Fibromyalgia Syndrome (FMS). This study aimed to characterise PCS related new-onset chronic MSK pain and its overlap with Fibromyalgia Syndrome (FMS).

**Methods:** Patients with new-onset chronic MSK pain following COVID-19 infection were enrolled and the nature of pain and associated symptoms captured using the C19-YRS (Yorkshire Rehabilitation Scale). FMS assessment was conducted as part of standard clinical examination using the American College of Rheumatology (ACR) 2010 criteria. Diagnosis of FMS was made when they meet the standard criteria of (1) Widespread Pain Index (WPI) ≥ 7 and Symptoms Severity (SS) score ≥ 5, or WPI is 3-6 and SS score ≥ 9, (2) symptoms have been present at a similar level for at least 3 months, and (3) the patient does not have a disorder that would otherwise explain the symptoms.

**Results:** Eighteen patients, twelve of whom were female, with an average age of 49.6 (SD 11.8) years and a Body Mass Index of 31.7 (SD 8.6) were enrolled. The average duration of symptoms from COVID-19 infection to assessment was 27.9 (SD 6.97) months. The new-onset chronic pain was widespread, primarily manifesting as muscle pain. Thirteen (72.2%) patients met the diagnostic criteria for FMS, with an average WPI score of 8.8 and an average SS score of 8.2, indicating a high level of pain and significant adverse impact on their quality of life.

**Conclusion:** The study found that 72.2% of the patients with new-onset chronic MSK pain following COVID-19 infection met the criteria for FMS. These findings support the hypothesis that FMS may develop as a long-term sequela of a viral infection, underscoring the need for further research into post-viral long-term conditions.

## 1. INTRODUCTION

Post-COVID syndrome (PCS), involves symptoms that persist beyond twelve weeks following an acute SARS-CoV-2 infection [1]. PCS is a novel multifaceted condition that presents a significant challenge to healthcare systems globally. It is estimated that a minimum of 10% of individuals who recover from COVID-19 develop PCS, with the prevalence increasing to 30% among moderate non-hospitalised cases and up to 70% in those who were hospitalised [2]. In the United Kingdom alone, approximately two million people are currently experiencing symptoms of PCS, highlighting its extensive impact [3]. One of the common and enduring symptoms of PCS is new-onset chronic Musculoskeletal (MSK) pain [4,5]. This adds a considerable strain to the already substantial global burden of chronic MSK pain, estimated to affect 1.7 billion individuals worldwide before the COVID-19 pandemic [6]. New-onset MSK pain in PCS is associated with a wide array of symptoms, including fatigue, sleep disturbances, and cognitive issues. These symptoms are similar to those observed in Fibromyalgia Syndrome (FMS) [7–9].

Fibromyalgia syndrome is a complex and chronic condition characterised by widespread pain, fatigue, sleep problems, and cognitive disturbance, which significantly impair the quality of life [10]. It affects between 2% and 8% of the general population, with a higher prevalence in females and middle-aged individuals [11]. FMS is widely recognized as being associated with central and peripheral nervous system sensitization [12,13]. This results in pain that often manifests as widespread aching and burning. FMS has been observed alongside other chronic pain conditions such as Osteoarthritis and Rheumatoid Arthritis. Notably, about 10% to 30% of patients with rheumatic disorders also meet the criteria of FMS, often referred to as secondary fibromyalgia [14]. Despite the high prevalence of FMS and its well-documented clinical characteristics, its precise aetiology remains largely unclear. A few studies have shed light on potential triggers for the development of FMS, pointing toward inflammatory and autoimmune disorders, as well as viral infections as possible contributing factors [15,16]. The association between FMS and specific infections has been studied, revealing links with various infection agents, including Epstein-Barr virus, Q fever, viral hepatitis, HIV, and Lyme disease [16,17].

Recent research has begun investigating the potential association and similarities between FMS and SARS-CoV-2 virus, expanding our understanding of how infections might trigger the development of FMS [8,9,18,19]. Studies have identified a notable overlap in the symptoms and pathophysiological mechanisms of PCS and FMS, suggesting that SARS-CoV-2 virus could play a pivotal role in mediating FMS. This emerging evidence points to a substantial occurrence of FMS among those recovering from COVID-19, underlining the need for further investigation into the relationship between infectious diseases and chronic pain syndromes.

The aim of this study was to characterise the nature of new-onset chronic MSK pain in individuals with PCS and explore its commonalities with FMS, utilising validated diagnostic tools and outcome measures. The research aimed to identify similarities in characteristics between PCS and FMS that could reveal underlying mechanisms and inform more effective treatment strategies for chronic MSK pain in individuals with PCS.

## 2. METHODS

### 2.1. Study design and participants

In this cross-sectional study, we assessed the characteristics of new-onset MSK pain and investigated its overlap with FMS. This data was derived from the Musculoskeletal Pain in Long COVID (MUSLOC) study, a prospective longitudinal study (clinicaltrials.gov NCT05358119). Participants eligible for this study met the following criteria: they were 18 years or older, had tested positive for COVID-19 or had COVID-19 symptoms confirmed by an independent clinician, received a clinical diagnosis of PCS according to the NICE guidelines, experienced new-onset MSK pain since their COVID-19 infection, were willing to adhere to study procedures, and could understand and read English. We excluded patients with pre-existing chronic MSK pain prior to their COVID-19 infection.

The Leeds Long COVID Community Rehabilitation Service acted as the primary recruitment site, supplemented by advertisement on social media, support groups, and research websites for wider outreach. Potential participants expressing interest were first contacted via telephone for eligibility screening. During this initial contact, they were fully briefed on the study’s requirements, including potential disadvantages, and benefits before providing their written informed consent. This research adheres to the ethical principles of the Declaration of Helsinki and was approved by London - Central Research Ethics Committee (REC), the Health Research Authority (HRA) and Health and Care Research Wales (HCRW) (Ref 21/PR/1377).

### 2.2. Procedure and outcomes

Demographic information including age, sex, ethnicity, marital status, body mass index (BMI), pre-existing co-morbidities, date of COVID-19 infection, hospitalisation due to COVID-19, number and dates of COVID-19 vaccinations, employment category and whether their employment status had been affected by COVID-19 pandemic and PCS were collected.

PCS associated symptoms were captured using the COVID-19 Yorkshire Rehabilitation Scale (C19-YRS) questionnaire [20]. It comprises a 0-10 Numerical Rating Scale (NRS) of main symptoms of PCS (including breathlessness, cough, swallowing, fatigue, continence, pain, cognition, post-traumatic stress disorder (PTSD), anxiety, depression, palpitations, dizziness, weakness, sleep problems, fever and skin rash) and their impact on five daily functions (including communication, mobility, personal-care, activities of daily living, and social role). Participants were also asked to grade their symptoms prior contracting COVID infection (pre-COVID). The score of items 1-10 adds up to the symptoms severity score (0-100), while items 11-15 scores add up to the functional disability score (0-50). The C19-YRS also includes a question capturing the overall health status before and after COVID infection (scores 0-10) in which a score of 0 means the WORST health the respondent can imagine and a score of 10 means the BEST health they can imagine.

The assessment of FMS was conducted as part of standard clinical examination in MUSLOC using the American College of Rheumatology (ACR) 2010 criteria [21]. It evaluates the Widespread Pain Index (WPI) by identifying 19 body areas where patients have experienced pain the previous week. Additionally, the ACR criteria examine Symptoms Severity (SS) across two parts: Part 1 quantifies the severity of fatigue, waking unrefreshed, and cognitive symptoms on a scale from 0 (no problem) to 3 (severe problems). Part 2 evaluates a broader spectrum of 40 other somatic symptoms, categorised as follows: no symptoms (score of 0), 1 to 10 symptoms (score of 1), 11 to 24 symptoms (score of 2), and 25 or more symptoms (score of 3) [22]. The criteria for diagnosing FMS are satisfied if all three of the following conditions are met: (1) WPI score of WPI ≥ 7 and SS score ≥ 5, or WPI is 3-6 and SS score ≥ 9, (2) symptoms have been present at a similar level for at least 3 months, and (3) the patient does not have a disorder that would otherwise explain the pain.

### 2.3. Statistical analysis

Statistical analysis was performed using IBM Statistical Package for the Social Sciences (SPSS) software version 29. Descriptive statistics were expressed as mean, standard deviation, minimum and maximum as appropriate for continuous variables, while numbers and frequencies were used for categorical variables.

## 3. RESULTS

### 3.1. Patient characteristics

A total of 18 patients were included in the study. The mean age of the patient was 49.6 (SD 11.8) years, comprising 12 females (66.7 %) and 6 (33.3 %) males. Body Mass Index (BMI) was 31.7 (SD 8.6). The mean duration of the onset of COVID-19 infection to the data collection point was 27.9 (SD 6.97) months.

### 3.2. Pain characteristics

Seventeen (94.4%) patients reported experiencing myalgia; of these, 15 (83.3%) were also experiencing arthralgia. Fourteen (77.8%) patients reported experiencing generalised widespread pain. The remaining patients, who did not report widespread pain, still experienced pain in at least four distinct body areas. Table 1 summarizes the demographic characteristics and distribution of general pain locations.

**Table 1.** Demographic and pain characteristics of the patients.

| Full sample (n=18) | n | % |
| --- | --- | --- |
| Age, years |  |  |
| <30 | 1 | 5.6 |
| 30-39 | 3 | 16.7 |
| 40-49 | 4 | 22.2 |
| 50-59 | 6 | 33.3 |
| 60+ | 4 | 22.2 |
| Sex |  |  |
| Female | 12 | 66.7 |
| Male | 6 | 33.3 |
| BMI (kg/m²) | Mean 31.7 (± 8.6) |  |
| Ethnicity |  |  |
| White | 14 | 77.8 |
| Asian | 4 | 22.2 |
| Black/ mixed/ other | 0 | 00.0 |
| Duration of LC symptoms (months) |  |  |
| <12 | 0 | 0.00 |
| 13-24 | 7 | 38.9 |
| 25-36 | 8 | 44.4 |
| 37-48 | 3 | 16.7 |
| +49 | 0 | 0 |
| **Number of COVID-19 infections** |  |  |
| Infected once | 4 | 22.2 |
| Infected twice | 10 | 55.6 |
| Infected 3 times | 3 | 16.7 |
| Infected 4 times | 1 | 5.6 |
| **Marital status** |  |  |
| Single | 4 | 22.2 |
| Married or in a domestic relationship | 14 | 77.8 |
| **Smoking status** |  |  |
| None | 17 | 94.4 |
| Active | 1 | 5.6 |
| **Comorbidities reported by patients (pre-existing before COVID-19 infection) \*** |  |  |
| Diabetes (type 1 or 2) | 1 | 5.6 |
| Hypertension | 2 | 11.1 |
| Chronic respiratory conditions | 3 | 16.7 |
| Chronic heart disease | 2 | 11.1 |
| Chronic neurological conditions | 2 | 11.1 |
| History of chronic musculoskeletal conditions | 2 | 11.1 |
| Chronic kidney disease | 0 | 0 |
| Chronic liver disease | 0 | 0 |
| Do not have any previous medical conditions | 6 | 33.3 |
| **Pain Frequency** |  |  |
| Intermittent (episodic) | 4 | 22.2 |
| Continuous | 14 | 77.8 |
| **Quality of pain\*** |  |  |
| Dull | 14 | 77.8 |
| Aching | 15 | 83.3 |
| Sharp and stabbing | 5 | 27.8 |
| Throbbing | 4 | 22.2 |
| Burning | 4 | 22.2 |
| Numbness | 3 | 16.7 |
| Crushing | 2 | 11.1 |
| Stinging | 0 | 0 |
| **Pain location reported by participants\*** |  |  |
| Widespread pain | 14 | 77.8 |
| Upper extremity | 11 | 61.1 |
| Lower extremity | 14 | 77.8 |
| Spine | 12 | 66.7 |
| MSK chest pain | 4 | 22.2 |

*\*Total percentages may exceed 100% as patients could report multiple responses.*

### 3.3. Results of C19-YRS

The data from C19-YRS showed a new-onset (de novo) of PCS symptoms compared to pre-COVID. All participants experienced pain and fatigue, while breathlessness was noted by 17 (94.4%) of them, predominantly during physically demanding activities such as stair climbing. Cognitive issues and anxiety were also prevalent symptoms, with being reported by 93% and 89% of the cohort, respectively. Additionally, fifteen patients (83.3%) reported having sleep problems.

There was also a significant increase in functional disability scores compared to levels before infection. PCS symptoms interfered with daily living activities for 17 (94.4%) of patients, impacting their ability to perform their family care and maintain social interactions. Many patients also reported detrimental effects on their communication abilities (15 patients), mobility (12 patients), and personal care (11 patients), underscoring a substantial burden to their daily lives compared to their pre-COVID status. Patients also observed a notable deterioration in their general health after contracting COVID-19, with overall health ratings decreasing from a pre-COVID average of 8.1 (SD 1.1) to 3.0 (SD 1.8) post-infection. The changes in symptom severity before and after COVID-19 infection are detailed in Table 2.

**Table 2.**
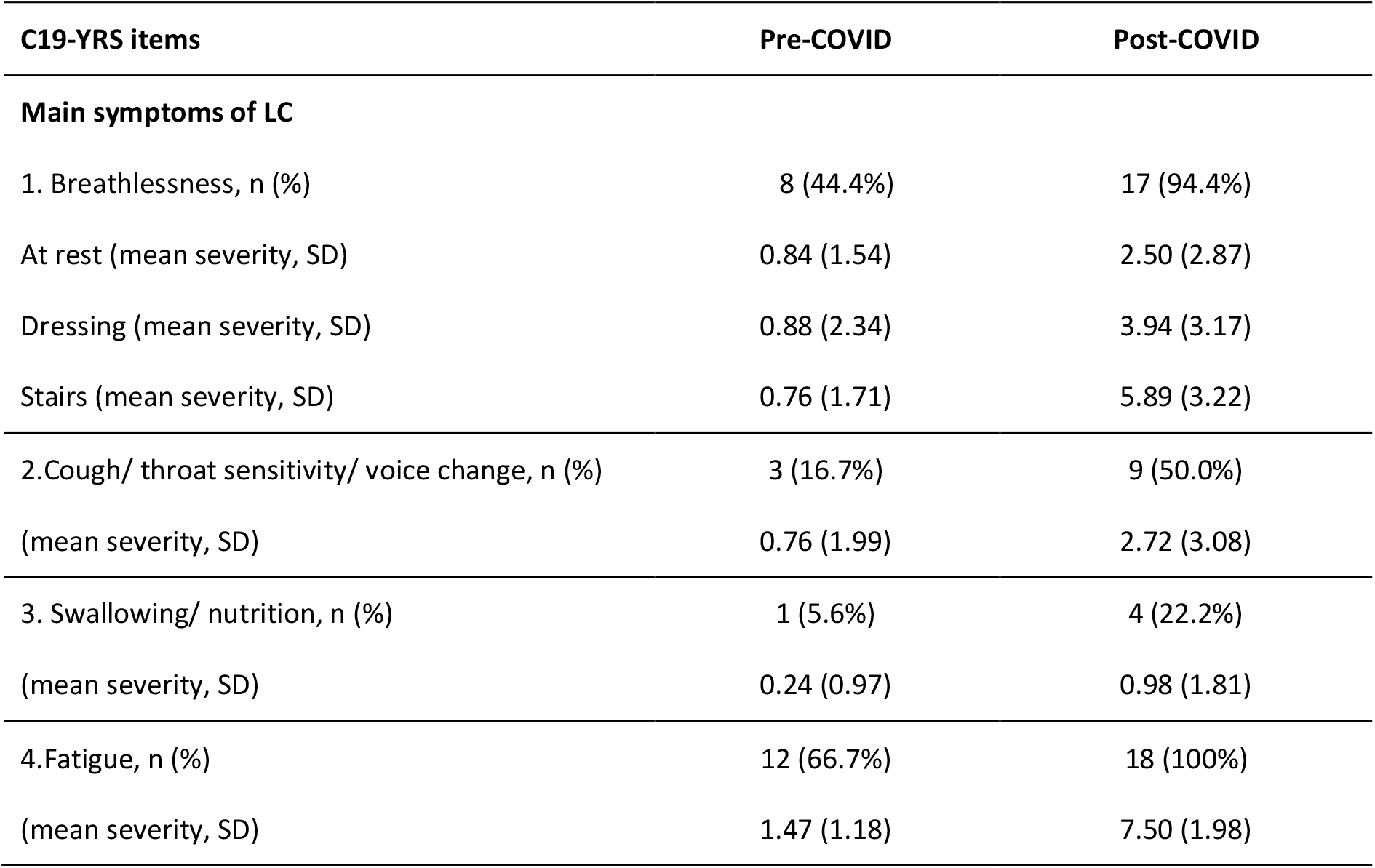
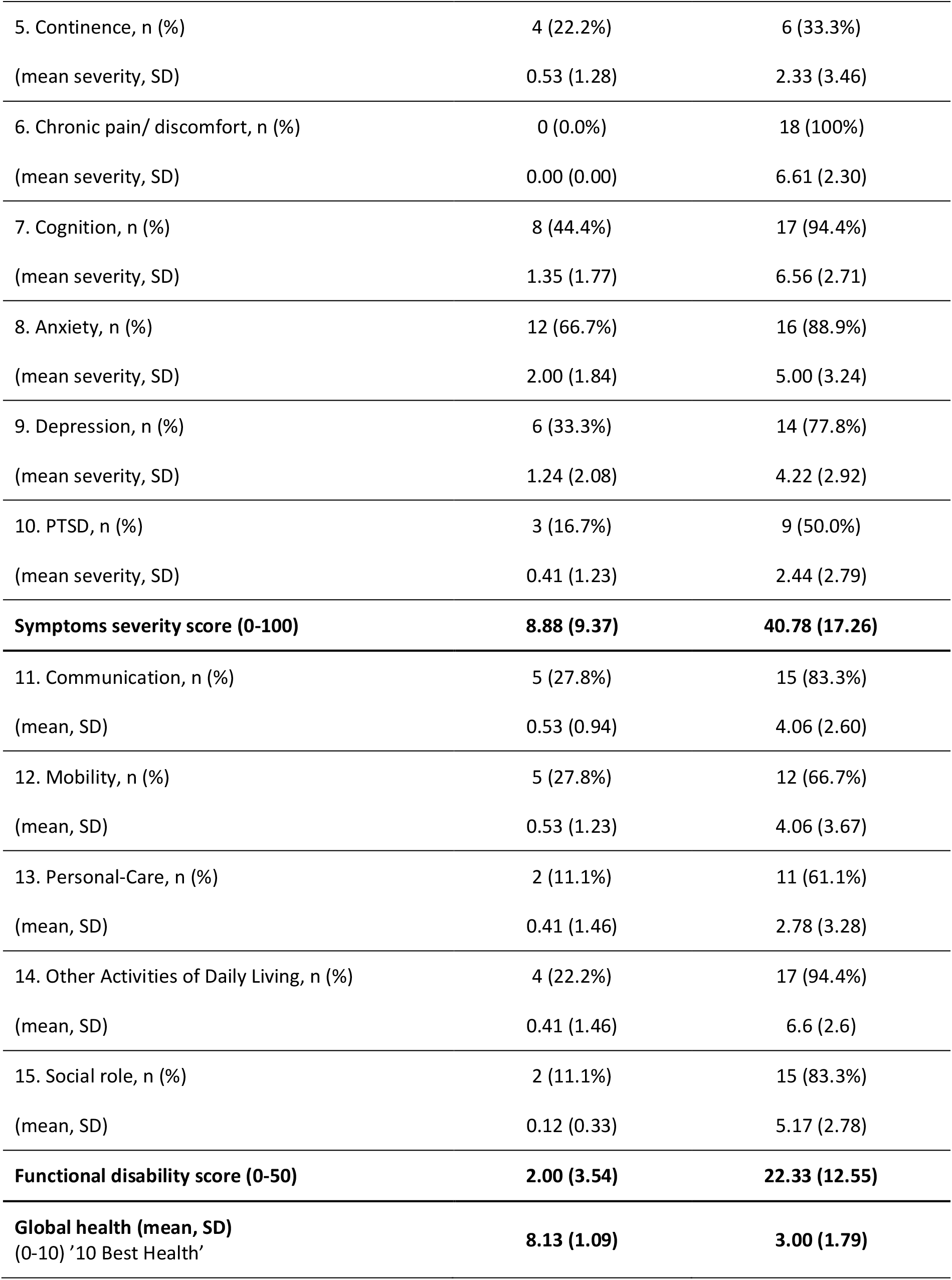
Summary of C19-YRS findings.

| C19-YRS items | Pre-COVID | Post-COVID |
| --- | --- | --- |
| **Main symptoms of LC** |  |  |
| 1. Breathlessness, n (%) | 8 (44.4%) | 17 (94.4%) |
| At rest (mean severity, SD) | 0.84 (1.54) | 2.50 (2.87) |
| Dressing (mean severity, SD) | 0.88 (2.34) | 3.94 (3.17) |
| Stairs (mean severity, SD) | 0.76 (1.71) | 5.89 (3.22) |
| 2.Cough/ throat sensitivity/ voice change, n (%) | 3 (16.7%) | 9 (50.0%) |
| (mean severity, SD) | 0.76 (1.99) | 2.72 (3.08) |
| 3. Swallowing/ nutrition, n (%) | 1 (5.6%) | 4 (22.2%) |
| (mean severity, SD) | 0.24 (0.97) | 0.98 (1.81) |
| 4.Fatigue, n (%) | 12 (66.7%) | 18 (100%) |
| (mean severity, SD) | 1.47 (1.18) | 7.50 (1.98) |
| 5. Continence, n (%) | 4 (22.2%) | 6 (33.3%) |
| (mean severity, SD) | 0.53 (1.28) | 2.33 (3.46) |
| 6. Chronic pain/ discomfort, n (%) | 0 (0.0%) | 18 (100%) |
| (mean severity, SD) | 0.00 (0.00) | 6.61 (2.30) |
| 7. Cognition, n (%) | 8 (44.4%) | 17 (94.4%) |
| (mean severity, SD) | 1.35 (1.77) | 6.56 (2.71) |
| 8. Anxiety, n (%) | 12 (66.7%) | 16 (88.9%) |
| (mean severity, SD) | 2.00 (1.84) | 5.00 (3.24) |
| 9. Depression, n (%) | 6 (33.3%) | 14 (77.8%) |
| (mean severity, SD) | 1.24 (2.08) | 4.22 (2.92) |
| 10. PTSD, n (%) | 3 (16.7%) | 9 (50.0%) |
| (mean severity, SD) | 0.41 (1.23) | 2.44 (2.79) |
| **Symptoms severity score (0-100)** | **8.88 (9.37)** | **40.78 (17.26)** |
| 11. Communication, n (%) | 5 (27.8%) | 15 (83.3%) |
| (mean, SD) | 0.53 (0.94) | 4.06 (2.60) |
| 12. Mobility, n (%) | 5 (27.8%) | 12 (66.7%) |
| (mean, SD) | 0.53 (1.23) | 4.06 (3.67) |
| 13. Personal-Care, n (%) | 2 (11.1%) | 11 (61.1%) |
| (mean, SD) | 0.41 (1.46) | 2.78 (3.28) |
| 14. Other Activities of Daily Living, n (%) | 4 (22.2%) | 17 (94.4%) |
| (mean, SD) | 0.41 (1.46) | 6.6 (2.6) |
| 15. Social role, n (%) | 2 (11.1%) | 15 (83.3%) |
| (mean, SD) | 0.12 (0.33) | 5.17 (2.78) |
| **Functional disability score (0-50)** | **2.00 (3.54)** | **22.33 (12.55)** |
| **Global health (mean, SD)**<br>(0-10) '10 Best Health' | **8.13 (1.09)** | **3.00 (1.79)** |

### 3.4. Results of ACR 2010 criteria for FMS

Thirteen (72.2%) of the evaluated patients, met the diagnostic criteria for FMS as defined by the ACR. The average WPI score among the patients was 8.8 (SD 5.1), indicating a high level of pain spread across multiple body regions. Additionally, the average SS score was 8.2 (SD 2.4), reflecting significant symptom severity related to fatigue, waking unrefreshed, cognitive symptoms, and the extent of other somatic symptoms. These scores collectively suggest a substantial impact on patients’ quality of life and align with the typical symptomology observed in FMS.

The remining five patients of the cohort, representing 27.8% did not meet the ACR criteria for FMS, with four of these five patients being males. This supports the evidence that FMS is more prevalent in females. The reasons for not meeting the ACR criteria varied and were related to their WPI and SS scores. specifically, one participant had a WPI of 8 but an insufficient SS of 4, failing to satisfy criterion 1a, indicating that while they experienced widespread pain, the severity of their symptoms was not adequately high. Two participants, one with a WPI of 5 and SS of 7 and another with a WPI of 5 and SS of 6, met the WPI required for criterion 1b, but their SS scores were too low, thus they did not meet the FMS criteria. Additionally, two participants with scores of WPI 0 and SS 3, and WPI 1 and SS 9 respectively, also failed to meet the criteria. The latter had a high SS score, indicating severe symptoms, yet the extremely low WPI made them ineligible for the FMS diagnostic criteria.

## 4. DISCUSSION

In this study, we explored the characteristics of new-onset MSK pain in individuals with PCS and its similarities with FMS. The research aimed to identify overlaps between PCS and FMS that could reveal underlying mechanisms and inform more effective strategies for both chronic MSK pain in individuals with PCS and in FMS. Our findings revealed that a substantial number of participants (72.2%) met the ACR criteria for FMS, underscoring a significant overlap between PCS and FMS. This demonstrates a high prevalence of widespread pain and symptomatology consistent with FMS in the PCS population.

The findings of this study align with and expand upon recent research exploring the connection between FMS and SARS-CoV-2 virus, enhancing our understanding of how viral infections may trigger chronic long-term conditions such as FMS. For instance, Haider et al. studied 203 PCS patients, identifying a subset whose symptoms closely resembles FMS, negatively impacting cognitive and physical function. Their data also revealed that 8.4% of those patients met the ACR criteria for FMS. Notably, the study also included another group of 22 patients who had already been diagnosed with both PCS and FMS, underscoring a significant overlap in the symptomatology of both conditions [9]. Additionally, Ursini et al, reported that 30.7% of individuals with PCS meet the FMS diagnostic criteria, suggesting a substantial prevalence of FMS manifestation among those who recovered from COVID-19 [18]. In another study, Savin et al. investigated the incidence of FMS in 198 patients who discharged after COVID-19 hospitalisation, it was found to be 15%, with a higher prevalence in females (63%) [23]. These are considerably lower than the 72.2% identified in our study, indicating varying levels of FMS manifestations across different cohorts and different criteria used for diagnosis. Furthermore, Fialho et al, highlighted the similarities in pathophysiological mechanisms and symptoms of FMS and PCS, suggesting a potential mediating role of the SARS-CoV-2 virus in the development of FMS [8]. These studies collectively highlight the complex relationship between PCS and FMS and suggest a potential viral contribution to the chronicity of MSK pain, aligning with our findings of significant symptom overlap and functional impairment in PCS, similar to FMS patterns.

Although the connection between SARS-CoV-2 virus and FMS is still under investigation, several hypothetical mechanistic connections may be suggested based on what we know about the pathogenesis of these two different conditions. There might be similar pathophysiological pathways involved, the virus affects both the central and peripheral nervous systems, potentially leading to typical FMS symptoms like pain and sensory disturbances either by direct tissue damage or systemic inflammatory responses [8,9,23]. Additionally, the chronic stress and psychological impacts of the pandemic, such as isolation and ongoing anxiety, can exacerbate central sensitisation seen in both conditions [24,25]. Finally, the intense inflammatory response, often referred to as a “cytokine storm,” which is triggered by SARS-CoV-2 virus, can lead to chronic inflammation that affects the nervous system. This sustained inflammatory state could potentially initiate or aggravate FMS symptoms, underscoring a plausible link between COVID-19 and new-onset or worsening of FMS [15,16].

This study has several limitations including the absence of a control group and reliance on patient reported questionnaires. We however need to acknowledge that these conditions are clinical syndromes and assessment is entirely reliant on subjective symptoms. The other limitation could be that those who did not meet the FMS criteria were assessed on a good day (we are aware that these are fluctuating conditions with good days and bad days). The relatively small patient group may also limit the generalisability of the findings. Despite these constraints, the study utilised validated diagnostic tools to rigorously assessing symptom overlap and highlights potential shared mechanisms, providing critical insights into our understanding of chronic widespread pain syndromes.

Future research should involve a larger sample size, have a control group (of those who had COVID-19 infection but no PCS symptoms) and follow both groups longitudinally with multiple assessment points (to incorporate the fluctuations seen in the conditions). We also need large mechanistic studies to better understanding the long-term impacts of persistent PCS and the development of FMS in individuals with PCS. Additionally, the growing body of evidence underscores the need for a deeper investigation into pathophysiological mechanisms of long-term syndromes after any infection, which could revolutionise approaches to diagnosis and treatment of post-infection sequelae.

## 5. CONCLUSION

Our study shows that new-onset chronic MSK pain following COVID-19 infection often meets the diagnostic criteria of FMS. These findings support the hypothesis that FMS can arise as a sequela of viral infections. Given these findings, it is critical to consider and screen for FMS in patients presenting with persistent PCS symptoms, particularly chronic pain and fatigue. This understanding not only challenges the current vagueness in validating PCS and FMS but also emphasises the need for further investment in clinical services and research to investigate the underlying mechanisms through which viral infections could lead to chronic conditions like FMS. The validation and management of FMS appropriately could potentially enhance patients’ understanding of the condition and the outcomes seen in services managing these conditions.

## Data Availability

All data produced in the present study are available upon reasonable request to the authors

## 6. ACKNOWLEDGMENTS

The authors would like to express their heartfelt gratitude to all participants and their families for their invaluable contribution to this study. Special thanks to the staff of the LCH Long COVID service for their crucial help in recruiting participants. We also acknowledge the support from the Leeds NIHR Biomedical Research Centre and Prof Paul Emery from the Leeds Institute of Rheumatic and Musculoskeletal Medicine.

